# Distributional challenges regarding data on death and incidences during the SARS-CoV-2 pandemic up to July 2020

**DOI:** 10.1101/2020.07.24.20161257

**Authors:** Kirsi Manz, Ulrich Mansmann

## Abstract

COVID-19 is a major global crisis with unpredictable consequences. Many scientists have struggled to make forecasts about its impact. Especially, appropriate preparations for a second wave are needed not to move in a costly panic mode again. It is necessary to get ideas about worst case scenarios regarding incidences, hospitalization, or use of ICU resources. They can be described in terms of extreme quantiles (95%, 99%, 99.9%) of specific distributions that supposedly formalize the data mechanism behind future observations.

Therefore, distributional issues do matter. Cirillo and Taleb argue that a natural and empirically correct framework for assessing and managing real risk in pandemics is provided by extreme value theory dealing with extrema and not averages. We explore this idea in more detail.

In this paper we discuss the fat-tail patterns in the distribution of the global COVID-19 data by analyzing data from 66 countries worldwide. We also explore their relevance at a lower, regional scale perspective (national, federal state), which is in our opinion more relevant for planning measures against the epidemic spread. For this we analyze data from the German federal state of Bavaria.

We conclude that fat-tail patterns are seen in global data, possibly reflecting the respective heterogeneity between different countries regarding incidences and fatalities during the ongoing epidemic. However, the disease activity at regional level seems to be better described by classical Poisson based models. To bridge the gap between regional and global phenomena we refer to mixtures of slim-tail distributions that may create fat-tail features.

Especially in the beginning of a pandemic acting according to the “better safe than sorry” principle and taking extreme forecasts as the basis for the decisions might be justified. However, as the pandemic continues and control measures are partially lifted, there is a need for a careful discussion how to choose relevant distributions and their respective quantiles for future resource planning in order not to cause more harm as the pandemic itself.

## Introduction

The SARS-CoV-2 pandemic stimulated multiple modeling groups to develop a rich toolbox of statistical, mathematical, and computational instruments. Their interest was to forecast disease trajectories, to assess effects of interventions, and to understand the dynamics of the epidemic^1–9^. The conceptual basis of these models builds on the methodological background within the communities to which the developers belong^7, 10, 11^. The corresponding background determines the horizon of their model building.

Some models became tools for policy and decision makers^12^. They were helpful in developing greater situational awareness regarding potential risk implied by the epidemic: What is the chance that the health care system breaks down, that capacities of ICU units are exhausted? The aim of using these models is to derive probabilities for bad case or worst case scenarios and to compare the effects of different interventions with respect to the predicted occurrence of worst cases. To this end it is necessary to think in distributions and not in terms of location parameters. Bad or worst case scenarios can be defined by high or extreme quantiles of the specific distributions like the distribution of maximal daily case load during a second wave of the COVID-19 epidemic.

Pasquale Cirillo and Nassim Nicholas Taleb recently published a paper^13^ in which they emphasize that a natural and empirically correct framework for assessing (and managing) the real risk of pandemics has to consider heavy tailed risks. They argue that extreme value theory (EVT) is an appropriate starting point when thinking of the damage a pandemic may create. They formulate what we would call a *distributional challenge*. Their ideas are illustrated by a historical data set on the distribution of deaths from the major epidemic and pandemic diseases of history, from 429 bc until the present. This is a very heterogeneous collection of data and there may be a heavy selection bias since the most famous outbreaks in human history are preferentially selected from the extreme tail of the distribution of all outbreaks^14^. Therefore, Cirillo and Taleb’s analysis does not convincingly show that heavy-tail distributions are behind the data of the ongoing pandemic. We analyzed current data to find out, whether the worldwide distribution of fatalities related to COVID-19 seems to follow a fat-tailed distribution.

One needs to bear in mind that there are different scales at which to look at the ongoing pandemic, a *global* and a *national* or a *regional* one. Cirillo and Taleb took a look at the data at the global level, which is per se heterogeneous. In most cases decision making is not done at the global but at the national or even the regional level, thus it is related to structures with less heterogeneity. We asked ourselves if fat-tail aspects are still present at these lower scales, where uncertainty and heterogeneity still exist but might be captured using corresponding distributional mixtures. Therefore, we explore regional data from Germany using Bayesian inference and Markov chain Monte Carlo (MCMC) strategies to combine uncertainty in estimation with uncertainty in prediction. We reflect on the tail properties of predictive distributions of interest for measures like peak incidences, total incidences, and days with a critical amount of incidences.

The goal of the present paper is to study distributional aspects of the actual pandemic and to provide an example to demonstrate the use of mixture distributions to end up with helpful ideas concerning bad or extreme case scenarios. We also emphasize the use of the Lorenz curve^15^ and the Gini index^16^ to visualize and quantify heterogeneity of distributions.

## Results

### Fat-tail assessment in the global death data

On June 30 2020 worldwide 169 countries have reported a total of 511,137 COVID-19 related deaths. Of them 25% have occurred in the US only (127,417/511,137). Sixty-six countries have reported at least 200 deaths in confirmed COVID-19 cases and were included in the analyses regarding heavy tails. Table S1 in Appendix reports the death counts as of June 30 2020 per country, population and rescaled death rate per 100 000 persons for countries with at least 200 reported deaths in persons with confirmed SARS-CoV-2 infection.

Figure 1 represents the exploration of the global data on COVID-19 fatalities. Panel A shows the histogram of the reported number of deaths for the 66 countries. The empirical mean (median) is 7671 (1385). The empirical standard deviation (SD) is 18648. Since standard deviation is not a reliable measure for possibly fat-tailed distributions, we also calculated the mean absolute deviation (MAD - mean of all absolute deviations of each data point from the overall mean): 9657. Empirical skewness and kurtosis are 4.5 and 24.0, respectively. They are based on the third and fourth moments of a distribution and the empirical estimates may not be quite reliable. Panel B shows the maximum to sum plot^17^ and further indicates that these estimates are problematic. Here the ratios of partial maximum 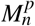 and partial sum 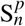 for the first four statistical moments *p* (*p*=1: mean; *p*=2: variance; *p*=3: skewness; *p*=4: kurtosis) are shown over the number of used observations *n*. Since no ratio converges towards zero, the plot suggests that no finite moment is likely to exist for the death counts. Therefore, it could be problematic to use the mean, standard deviation, skewness, or the kurtosis derived from the data. Panel C shows the Zipf plot^17^, where the empirical survival over the reported deaths is plotted. To guide the eye, a simple linear fit to the data is shown as a red line. Panel D shows the Hill plot^17^ with an estimate of *ξ*. For the definition of *ξ* we refer to the Methods section and Appendix B. It seems that *ξ* may be above 1, supporting the claim of heavy tails. Panel E shows the quantile-quantile plot (QQ plot) supporting our suggestion of infinite moments for the COVID-19 fatalities. The Lorenz curve is shown in Panel F, one for the distribution of cumulative incidences between countries (blue) and a second one for the distribution of cumulative fatalities between countries (red). The corresponding Gini indices are 0.86 (incidences) and 0.91 (fatalities).

**Figure 1.**
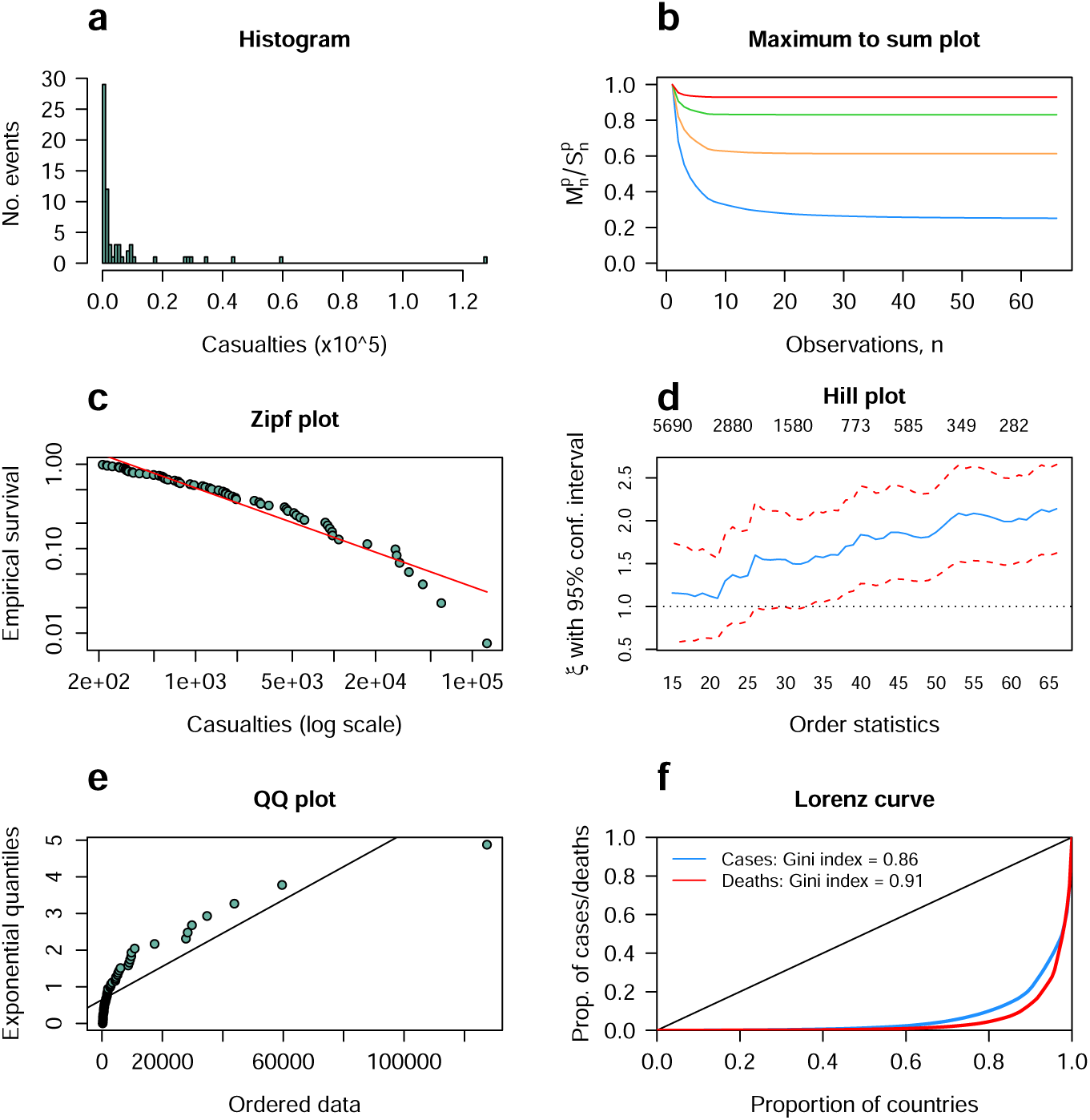
Exploration of fat-tail features of the global COVID-19 fatality data. All countries with at least 200 reported COVID-19 related deaths were included.

We repeated the analyses using data from all countries with 1, 500 and 1000 reported deaths as the minimum, respectively. Qualitatively similar results as for our cutoff of 200 deaths were obtained (data and code available from https://www.en.ibe.med.uni-muenchen.de/research/heavy-tail-issues/index.html).

### Bayesian inference and prediction of future incidences: A Bavarian case study

After a locally controlled COVID-19 outbreak in January 2020, Bavaria faced a federal state-wide outbreak after the winter vacation 2020, starting end of February. Daily data on incidences and fatalities is available from 27th of February to 30th of June 2020. During this time a total of 48366 confirmed incidences and 2580 deaths were reported. We use a standard Bayesian prediction approach which combines the posterior distribution of the model parameters with the uncertainty of future events. The events will be alternatively analyzed using Poisson as well as the negative binomial (NB) distribution. The corresponding parameters are their log-transformed means over time. In case of the NB distribution we assume p=0.1 (corresponding to variance being 10 times larger than the mean) to put more weight into the long tail of the distribution.

Panel A in Figure 2 shows the time course of the observed incidences on a log-transformed scale (plotted by black dots). Since the log-transformed incidences of cases look close to linear during the increasing and the decreasing phase of the epidemic for simplicity we decided to model the time course by a linear change point function: *α* + *β*_1_ · (*t*− *γ*) ·1_[*t<γ*]_ + *β*_2_ · (*t*− *γ*) ·1_[*t*≥ *γ*]_. Panel A also shows the posterior distributions of the linear change point model representing the log-transformed mean time course assuming Poisson (red lines) and negative binomially (grey lines) distributed observations. Since the NB model assumes larger variance the corresponding mean is expected to be lower compared to the Poisson model. Panel B represents predictions for future epidemics which follow the same pattern as infered from the present observed episode. The predictive distribution under the Poisson model (represented in red) is more focused than the one under the NB model (represented in grey). Panel C provides a description of the predictive peak number distribution of a future epidemic under both modelling strategies. The horizontal dashed lines show the six highest daily incidences seen during the current episode. In median, both models predict a similar number of peak incidences, which is slightly below the observed highest daily incidence. Panel D shows the predictive distribution of cumulative incidences over a period of 125 days of a future epidemic together with the actually observed cumulative number of incidences (horizontal dashed line). The NB model predicts a lower cumulative number of future incidences than the Poisson model does. In median, the Poisson model predicts a comparable number of cumulative incidences as was observed in the current data.

**Figure 2.**
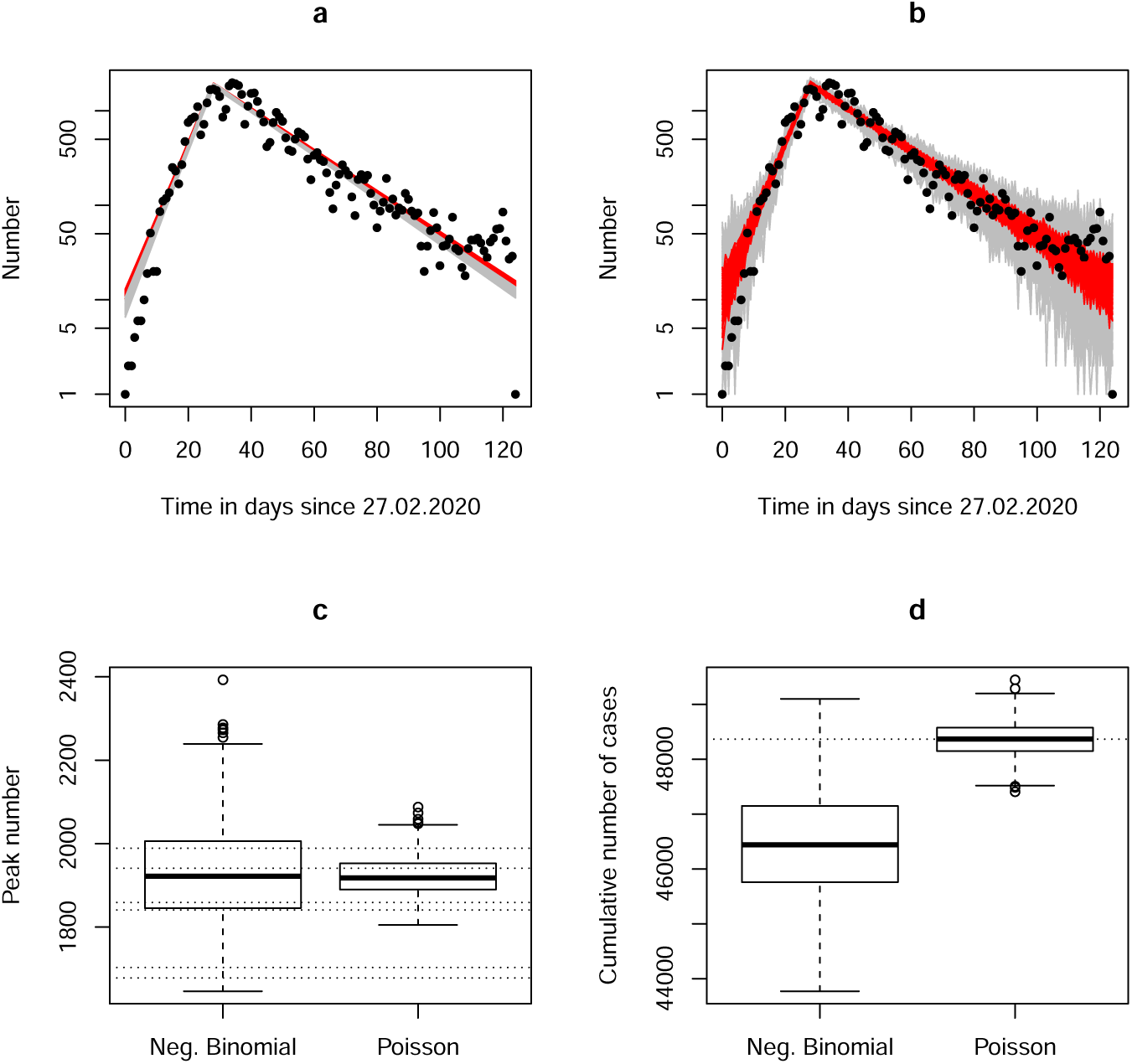
Results of the Bayesian analysis for Bavarian incidences.

Similarly, we determine the distributions of *critical days* of a future epidemic. We define a *critical day* as a day with more than 1000 incidences. Table 1 summarizes the results of 1000 samples from the posterior distribution of *critical days*. For the definition of worst cases we look at the quantiles of the predictive distributions of peak incidences or cumulative incidences during the epidemic, as shown in Table 2.

**Table 1.** 1000 samples from the posterior of *critical* days. NB = negative binomial.

| Distribution | 12 days | 13 days | 14 days | 15 days | 16 days | 17 days | 18 days | 19 days |
| --- | --- | --- | --- | --- | --- | --- | --- | --- |
| Poisson | 0 | 0 | 0 | 24 | 457 | 470 | 49 | 0 |
| NB | 2 | 32 | 173 | 288 | 311 | 152 | 40 | 6 |

**Table 2.** 1000 samples from the posterior of peak incidences and cumulative incidences. NB = negative binomial.

| Outcome | Distribution | 50% Quantile | 80% Quantile | 95% Quantile | 99% Quantile |
| --- | --- | --- | --- | --- | --- |
| Peak Incidence | Poisson | 1918 | 1960 | 2000 | 2039 |
| Peak Incidence | NB | 1922 | 2025 | 2120 | 2224 |
| Cum. Incidence | Poisson | 48370 | 48630 | 48840 | 49060 |
| Cum. Incidence | NB | 46440 | 47282 | 47990 | 48560 |

A linear change point function was also fitted to the logarithm of observed fatalities in Bavaria. As in case of incidences, a Poisson model and a NB model with p=0.1 were the respective counting distributions. The corresponding results are shown in Figure 3. Panel A shows the time course of the observed deaths between February 27th and June 30th, 2020, on a log-transformed scale. The observed daily fatalities are represented by black dots and the posterior distributions of the linear change point model assuming Poisson and negative binomially distributed observations as red and grey lines, respectively. Panel B represents predictions for future epidemics assuming the same pattern as infered from the present observed episode. As in case of incidences, the predictive distribution under the Poisson model (represented in red) is more focused than the one under the NB model (represented in grey). Here, the NB model leads to very broad prediction intervals. Panel C provides a description of the predictive peak number distribution of a future epidemic under both modelling strategies. The horizontal dashed line shows the highest daily death count seen during the current episode. The peak death counts predicted by the NB model are higher than for the Poisson model and both models predict in median higher peak death counts as observed in the current data. Panel D shows the predictive distribution of cumulative number of deaths over a period of 125 days of a future epidemic together with the actually observed cumulative number of deaths, as represented by the horizontal dashed line.

**Figure 3.**
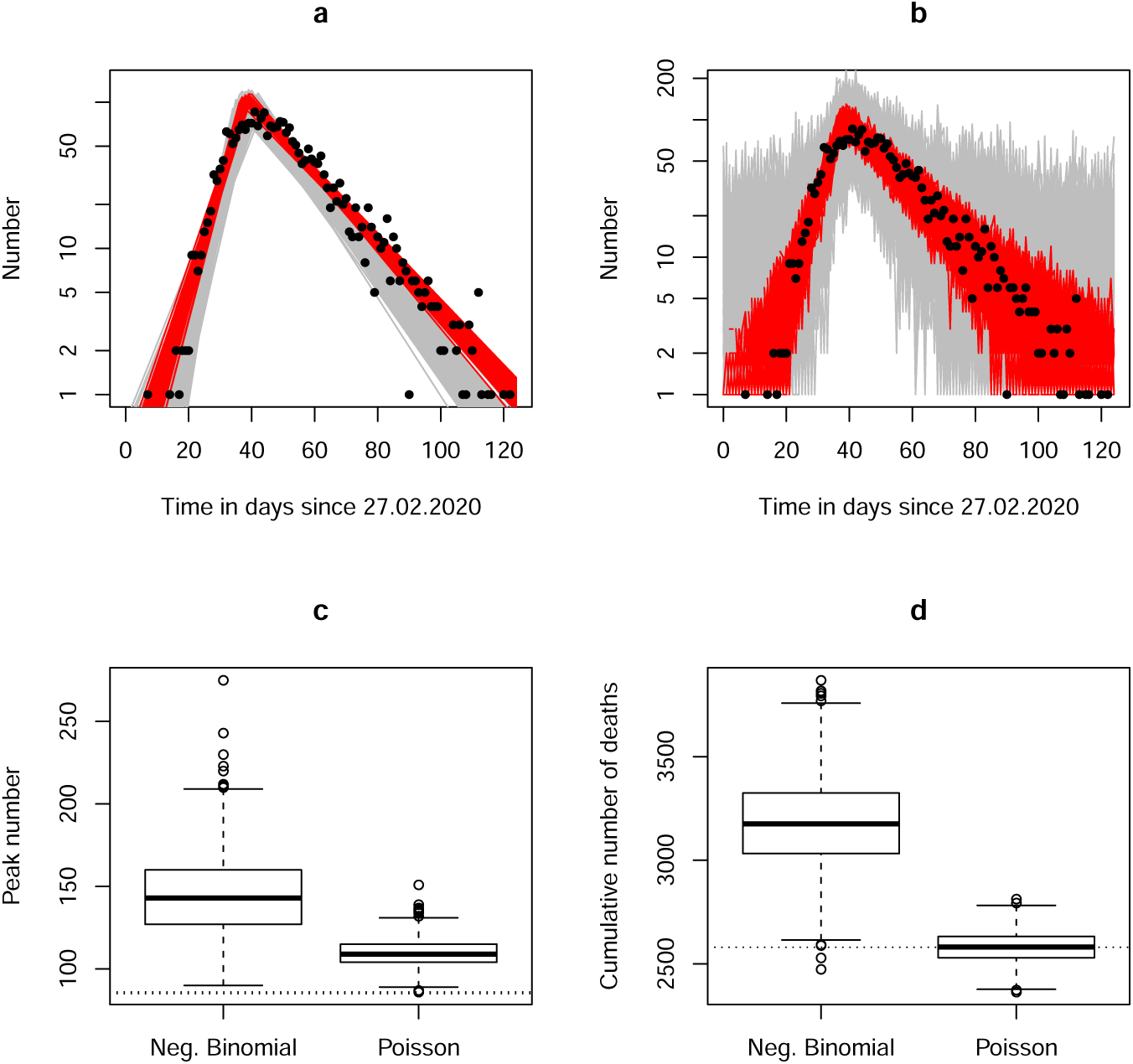
Results of the Bayesian analysis for Bavarian fatalities.

The Bayesian analysis using Poisson or NB counting models (with a large overdispersion) results in nearly symmetrical predictive distributions of relevant pandemic parameters. In both examples (three months of Bavarian incidences and deaths) the Poisson distribution may even provide a more reliable approach to prediction compared to the NB model. The conclusion is simply that heavy-tail or fat-tail features do not appear to be relevant when modelling the epidemic at a regional level.

The details of the Bayesian approach are given in Appendix C, where we also provide the data within a WinBUGS doodle and an R script for reproducing the results.

## Discussion

This paper explores the claim of Cirillo and Taleb^13^ that tail risks of epidemics have to be taken into account in order to develop appropriate planning and decision strategies. Our findings are simple: On the global level, when investigating COVID-19 incidence and death data across nations worldwide we find the heavy-tailed patterns Cirillo and Taleb were concerned about. However, these properties disappear when one moves to the level of single nations and specific regions where decision making and planning is mostly performed. Therefore, the warnings of Cirillo and Taleb may not apply to the respective national or federal administrative perspectives.

Globally the distribution of COVID-19 fatalities seems to follow a fat-tailed distribution. Our empirical analysis points towards *ξ* above 1 that proposes a tail exponent in the heavist class^13^. This seems to support the claim that pandemics are extremely fat tailed. Some countries only report few cases, whereas another countries such as the USA are heavily hit by the pandemic. Concerning the incidence data we found that 10% of the countries (13 in total) are responsible for 80% of all reported cases. For deaths the unequality is even more pronounced, since only 7% of the countries (13 countries) are responsible for 80% of all reported deaths.

Our heavy tail analysis uses truncated data, since only countries with at least 200 documented deaths were taken into account. Nevertheless, the observation of the heavy-tailedness of the worldwide distribution of the respective data seems to hold for any minimal number of considered deaths, which is a characteristic of the scaling properties (i.e. self-similarity) of power laws. The heavily affected countries are the ones appearing in the tail of the distribution and not the countries with only few infections. Thus lowering the death count cutoff and adding more observations into the “bulk” of the distribution function would not let the tail disappear. Even if the historical data set used by Cirillo and Taleb might only contain the most famous reported outbreaks and suffer from selection bias, the heavy tailedness would still probably hold, if they would have had information regarding other not so prominent smaller outbreaks, too. Our analysis avoids the sampling bias commented by Ioannidis et al.^14^ by looking at the complete data set of the global COVID-19 epidemic. However, there is a natural upper boundary in the data, since no pandemic can kill more people than the current world population. Cirillo and Taleb performed empirical exploration of these upper boundary effects on the heavy-tail properties.

Cirillo and Taleb^13^ provide several simple statistical tools to explore the heavy-tail property of the distribution behind empirical data. Besides heavy tailedness also the heterogeneity between countries is of great interest to monitor. We propose to add the Lorenz curve^15^ and the Gini index^16^ to this toolbox. The Lorenz curve visualizes and the Gini index quantifies heterogeneity within a distribution. For both the incidences and the deaths a high Gini index was found. This heterogeneity between countries can be a consequence of differences in diverse infrastructures, such as the reporting system, the health care system, or the effectivity of implemented control measures.

The message of Cirillo and Taleb is that forecasting single variable in fat-tailed domains violates common sense and probability theory^18^. They claim that risk management is concerned with tail properties and distributions of extrema. According to our observations the fat-tail property is only present at a global level and disappears at a lower scale (nation or region). We explore this issue for the federal state of Bavaria with a population of about 13.08 million people by looking at the epidemic from end of February 2020 to end of June 2020. It is our interest to apply Bayesian inference and prediction techniques to explore their usefullness.

Bayesian methodology allows to take into account the uncertainty regarding the main parameters of the model and to combine posterior predictions of parameters with relevant distributions to make predictions. We provide a Bayesian toy example to illustrate how to accomplish the analysis of tail properties and distributions of extrema by looking at the 90%, 95% or 99% quantiles of the respective predictive distributions. We are interested in the distribution of the peak incidences, the number of critical days (with more than 1000 incidences), and the total number of events during a three months periode. By following straightforward Bayesian arguments we build our models on two distributional assumptions: the Poisson distribution as a workhorse for epidemiological incidence models and the NB distribution as a candidate for massive overdisperion.

A characteristic of the Poisson distribution is that its mean and variance are equal. To capture more variation in the data the NB model can be used. It includes a parameter for dispersion and is thus suited for situations where the variance is markedly larger than the mean. In our modelling approach we allowed the variance to be 10 times the mean (corresponding to p=0.1). We found out that the NB model tended to underestimate future number of cumulative incidences and overestimate future number of cumulative fatalities, as shown in panel D of Figures 2 and 3. The Poisson model on the other hand predicted in median a comparable number of incidences and fatalities for a future episode as the actual data showed. For both incidences and deaths we would prefer the Poisson model over the NB model to derive critical quantiles of predictive distributions. Allowing for a large variance seemed not to be helpful in our example and could be a critical issue when deciding on modelling strategies for prediction.

There are theoretical concepts which may explain the qualitative changes in distributional features between regional and global levels of the epidemic. The mixture of distributions without heavy tails can result in a distribution with heavy tails. For example, the Pareto distribution, very common in EVT, can be derived as a gamma mixture of exponential distributions. It is a power-law distribution that is used in description of social, scientific, geophysical, actuarial, and many other types of observable phenomena. The mixture resulting in the Pareto distribution is a special case of the gamma-gamma distribution presented in the Methods section. Venturini et al.^19^ apply this concept in health economy. Typically, medical expenditures present highly skewed, heavy-tailed distributions. In general, simple variable transformation is insufficient to achieve a tractable low-dimensional parametric form and nonparametric methods do not exists for efficiently estimating exceedance probabilities for large thresholds. Motivated by this context, they propose a general Bayesian approach for the estimation of tail probabilities of heavy-tailed distributions, based on a mixture of gamma distributions in which the mixing occurs over the shape parameter.

The analyses we showed for Bavaria should be understood as a demonstration of the methods of how one might analyze data in preparation for the second wave. For simplicity we ignore many aspects of a trustworthy model. For example there is no correction for underreporting or adjustment to demography of the population. Our model is weak regarding the assessment of the effects of specific interventions, too. This could be only reflected in the slope of the decreasing part of the change point model. The chosen change point model allows to fit two exponential functions to the data, one for the increasing part of the curve up to maximum and one for the decreasing part after the peak. Additionally, the change point itself is the third parameter. There are examples where modellers have applied alternative three-parameter models to the overall time course of the epidemic by assuming it follows a scaled Gaussian density^10^ or scaled gamma density. Our experience with the scaled gamma density showed a poor fit to the observed time course. We made similar observations as Buchanan commented^20^: These models put too severe restrictions on the up and down of the observations. Especially, the Gaussian assumption implies that the incidences follow a bell-shaped curve. This assumption guarantees that the pandemic disappears as fast as it developed.

Additionally, for the prediction we assumed that the second wave would follow the same pattern as the current episode does. This may be a conservative assumption, since we have hopefully learned from the first wave and would react more appropriately next time.

Our analysis suffers from limitations regarding the data quality, of which underreporting of cases and deaths might be the most prominent one. We do not apply nowcasting to correct for reporting artefacts in the data^21^. We also do not consider testing strategies to adjust incidence reporting. Our model does not allow to implement complex intervention strategies. We simply assume that the next wave follows the same dynamics as the first wave and ignore alternative approaches like analysing excess deaths during the epidemic^22^. Therefore, the predictions we are providing are illustrative and may not reflect future scenarios. It is not our goal to provide a new prediction model but to emphasize that MCMC based Bayesian prediction is a very suitable tool to address to probability of extreme outcomes.

To conclude, in this paper we investigated both global and regional COVID-19 data. We found fat-tail properties in the global data but within a country or even smaller scale of federal states we believe that this phenomenon is currently not seen. The Gini index provides an additional tool to investigate heterogeneity in incidences and deaths between countries, which was found to be high. We showed an example for a prediction of a second wave using a Poisson based model combined with Bayesian methods, where we emphasized to take a look at the extreme quantiles of the predictive distribution and the duration of time, during which a critical amount of incidences or deaths might be reported. Our discussion might motivate the use of Bayesian strategies for planning of future healthcare resources.

## Methods

### The Data

Two data sets are used: Global data from the world population and regional data from the federal state of Bavaria (Germany). The global death and incidence data was retrieved from the COVID-19 Data Repository by the Center for Systems Science and Engineering (CSSE) at Johns Hopkins University^23^. We excluded any cruise ship data. The population data for each country was taken from the European Centre for Disease Control and Prevention (ECDC) COVID-19 database^24^, where a population estimate for the year 2019 is provided. The data is shown in Supplementary Table S1. The regional Bavarian data was provided by the Bavarian Health and Food Safety Authority (LGL). It contains information on the daily number of reported infections and reported deaths up to June 30 2020 for all 96 Bavarian districts. The relevant data used for the modelling is provided in the Supplementary Table S2.

The data were analysed using R Version 3.6.3^25^ and WinBUGS Version 1.4.3^26^.

### Fat-tail distributions

A non-negative continuous random variable *X* is *fat tailed* if its survival function *S*(*x*) = *P*(*X* ≥ *x*) decays as a power law 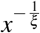 the more one moves into the tail. For example, the survival function of a Pareto distribution is (*x*_*m*_*/x*)^*α*^ for *x > x*_*m*_ *>* 0 and a positive shape parameter *α*. That is 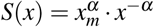 and 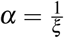. The parameter *ξ* > 0 is known as the tail parameter, and it governs the ‘fatness’ of the tail: the larger *ξ*, the fatter the tail. Moreover, *ξ* determines the existence of moments. The moment of order p exists, that is *E*[*X* ^*p*^] *<* ∞, if and only if 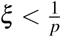. This implies that the *p*th moment of a Pareto distribution does exist if *α > p*. For *α*≤ 1, no moments exist at all. Cirillo and Taleb^13^ provide different graphical analysis tools to study the fat-tail property of a distribution: *maximum-to-sum plot, meplot, Zipf plot*, and *Hill plot*. The quantile-quantile (QQ) plot of the ordered data against the exponential distribution offers another tool to investigate if the data could be fat tailed. As suggested by Embrechts^17^ and applied by Cirillo and Taleb^27^, a QQ plot showing concave behaviour supports the claim of a fat-tailed distribution. More technical details are given in Appendix B.

It is also important to note that fat-tail distributions can arise from mixtures of non-fat tail distributions. An example is the Gamma-Gamma distribution (GGd) as discussed on page 337 of Held and Bové^28^. The GGd GG(*α, β, δ*) has the parameters *α, β, δ >* 0 and the density

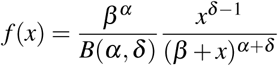

The function *B*(*α, δ*) is the beta function. The distribution GG(*α, β, δ*) is the mixture of the gamma distribution G(*δ, ε*) with shape parameter *δ* and rate parameter *ε*, where the rate parameter varies over G(*α, β*). Setting *δ* = 1 creates a gamma mixture of exponential distributions which results in a Pareto distribution (GG(*α, β*, 1)). Given the density of the GGd with *δ* =1 the result regarding the Pareto survival function (as shown in the second line of this section) follows by setting *β* = *x*_*m*_ and *α* = *α*.

### The Gini index of inequality and the Lorenz curve

The *Gini* index is widely used to characterize inequality in wealth and income^16^. The Gini index also quantifies inequality in the distribution. We do apply it to incidences and death counts attributable to COVID-19. The Gini index ranges from zero to one, where the value of zero would mean that the incidences or deaths are evenly distributed among the affected countries. On the other hand, the value of one would mean that all burden of COVID-19 incidences or fatalities lies upon one single country.

The *Lorenz* curve is closely related to the Gini index and serves as a visualization of inequality within a distribution^15^. In our application, the proportion of affected countries is shown in the horizontal axis and the proportion of all COVID-19 deaths is shown in the vertical axis. A diagonal line presents the line of equality, along which x% of countries are responsible for x% of the fatalities (corresponding to Gini index of zero). The Gini index is equal to the ratio of the area between curve and the area below the diagonal.

### Distributional mixtures, Bayesian inference and prediction

Bayesian inference based on MCMC methods allows to implement mixture distributions as posterior distributions for the parameters of interest. It is straightforward to sample from such distributions. This allows to address questions like: What is the (1 − *α*)% quantile for a specific parameter (determining worst or bad case scenarios)? To this end we apply a simple Bayesian MCMC procedure to the incidence and death time course data in Bavaria (Germany). The mean time course is parametrized by a triangle function for the log-transformed incidences: *f*_*α,β,γ*_ (*t*) = *α* + *β*_1_· (*t* − *γ*) · 1_[*t<γ*]_ + *β*_2_ · (*t* − *γ*) ·1_[*t* ≥ *γ*]_. This function breaks the symmetry between increase and decrease of the case numbers during an epidemic. The count data per day is modelled by a Poisson as well as a NB distribution. A MCMC machinery (WinBUGS) is used to sample future epidemic time courses (which are assumed to resemble the first wave). Based on these data and a run-in of 10000 cycles we derive the predictive distribution of peak epidemic incidences, of the number of days with more than 1000 incidences, and of the cumulative number of incidences using additional 1000 cycles. Details are given in Appendix C.

## Data Availability

All data generated or analysed during this study are included in this published article (and its Supplementary Information files).

## Data Availability

https://www.en.ibe.med.uni-muenchen.de/research/heavy-tail-issues/index.html

## Acknowledgements

We would like to thank Dr. Katharina Katz from the Bavarian Health and Food Safety Authority (LGL) for providing us the Bavarian data.

## Author contributions

U.M. conceptualized the idea. K.M. acquired and prepared the data. K.M. and U.M. analyzed and interpreted the data, drafted the manuscript and approved the submitted version.

## Competing interests

The authors declare no competing interests.

## Supplementary Information

## Appendix A the data sets

The raw data for the global data set was processed as follows: We extracted the total death count and the daily death counts per country between January 22 2020 and June 30 2020. For the Netherlands, United Kingdom and France we excluded the data from the following oversea territories (Netherlands: Aruba, Curacao, Sint Maarten, Bonaire, Sint Eustatius and Saba; UK: Bermuda, Cayman Islands, Gibraltar, Montserrat, Anguilla, British Virgin Islands, Turks and Caicos Islands and Falkland Islands (Malvinas); France: French Guiana, French Polynesia, Guadeloupe, Mayotte, New Caledonia, Reunion, Saint Barthelemy, St Martin, Martinique, Saint Pierre and Miquelon). In case of negative daily counts occuring as a consequence of data corrections we removed such data point(s) from the model fitting. Cumulative death data included all values and thus reflects the true number of reported deaths. For the analysis of heavy-tails we included data from all countries with at least 200 reported deaths in individuals with confirmed Sars-CoV-2 infection. The data is shown in Supplementary Table S1.

**Table S1.** Reported deaths as of June 30 2020 per country, population and rescaled death rate per 100 000 persons for countries with at least 200 reported deaths in persons with confirmed SARS-CoV-2 infection.

| No. | Country | Deaths | Population | Deaths per 100000 | No. | Country | Deaths | Population | Deaths per 100000 |
| --- | --- | --- | --- | --- | --- | --- | --- | --- | --- |
| 1 | US | 127417 | 329064917 | 38.7209 | 34 | Argentina | 1307 | 44780675 | 2.9187 |
| 2 | Brazil | 59594 | 211049519 | 28.2370 | 35 | Philippines | 1266 | 108116622 | 1.1710 |
| 3 | United Kingdom | 43801 | 66647112 | 65.7208 | 36 | Ukraine | 1173 | 43993643 | 2.6663 |
| 4 | Italy | 34767 | 60359546 | 57.5998 | 37 | Bolivia | 1123 | 11513102 | 9.7541 |
| 5 | France | 29763 | 67012883 | 44.4138 | 38 | Japan | 972 | 126860299 | 0.7662 |
| 6 | Spain | 28355 | 46937060 | 60.4107 | 39 | Algeria | 912 | 43053054 | 2.1183 |
| 7 | Mexico | 27769 | 127575529 | 21.7667 | 40 | Guatemala | 773 | 17581476 | 4.3967 |
| 8 | India | 17400 | 1366417756 | 1.2734 | 41 | Dominican Republic | 747 | 10738957 | 6.9560 |
| 9 | Iran | 10817 | 82913893 | 13.0461 | 42 | Afghanistan | 746 | 38041757 | 1.9610 |
| 10 | Belgium | 9747 | 11455519 | 85.0856 | 43 | Austria | 705 | 8858775 | 7.9582 |
| 11 | Peru | 9677 | 32510462 | 29.7658 | 44 | Panama | 631 | 4246440 | 14.8595 |
| 12 | Russia | 9306 | 145872260 | 6.3796 | 45 | Denmark | 605 | 5806081 | 10.4201 |
| 13 | Germany | 8990 | 83019213 | 10.8288 | 46 | Nigeria | 590 | 200963603 | 0.2936 |
| 14 | Canada | 8650 | 37411038 | 23.1215 | 47 | Hungary | 585 | 9772756 | 5.9860 |
| 15 | Netherlands | 6113 | 17282163 | 35.3717 | 48 | Sudan | 572 | 42813237 | 1.3360 |
| 16 | Chile | 5688 | 18952035 | 30.0126 | 49 | Moldova | 545 | 4043258 | 13.4792 |
| 17 | Sweden | 5333 | 10230185 | 52.1300 | 50 | Honduras | 497 | 9746115 | 5.0995 |
| 18 | Turkey | 5131 | 82003882 | 6.2570 | 51 | Armenia | 443 | 2957728 | 14.9777 |
| 19 | China | 4641 | 1433783692 | 0.3237 | 52 | Belarus | 392 | 9452409 | 4.1471 |
| 20 | Ecuador | 4527 | 17373657 | 26.0567 | 53 | Kuwait | 354 | 4207077 | 8.4144 |
| 21 | Pakistan | 4395 | 216565317 | 2.0294 | 54 | Czechia | 349 | 10649800 | 3.2771 |
| 22 | Colombia | 3376 | 50339443 | 6.7065 | 55 | Finland | 328 | 5517919 | 5.9443 |
| 23 | Egypt | 2953 | 100388076 | 2.9416 | 56 | Israel | 320 | 8519373 | 3.7561 |
| 24 | Indonesia | 2876 | 270625567 | 1.0627 | 57 | United Arab Emirates | 315 | 9770526 | 3.2240 |
| 25 | South Africa | 2657 | 58558267 | 4.5374 | 58 | Cameroon | 313 | 25876387 | 1.2096 |
| 26 | Switzerland | 1963 | 8544527 | 22.9738 | 59 | Yemen | 312 | 29161922 | 1.0699 |
| 27 | Iraq | 1943 | 39309789 | 4.9428 | 60 | North Macedonia | 302 | 2083458 | 14.4951 |
| 28 | Bangladesh | 1847 | 163046173 | 1.1328 | 61 | Korea, South | 282 | 51225321 | 0.5505 |
| 29 | Ireland | 1736 | 4904240 | 35.3979 | 62 | Serbia | 277 | 6963764 | 3.9777 |
| 30 | Romania | 1651 | 19414458 | 8.5040 | 63 | Norway | 250 | 5328212 | 4.6920 |
| 31 | Saudi Arabia | 1649 | 34268529 | 4.8120 | 64 | Bulgaria | 230 | 7000039 | 3.2857 |
| 32 | Portugal | 1576 | 10276617 | 15.3358 | 65 | Morocco | 228 | 36471766 | 0.6251 |
| 33 | Poland | 1463 | 37972812 | 3.8528 | 66 | Azerbaijan | 213 | 10047719 | 2.1199 |

Further regional German data for the federal state Bavaria, provided by the Bavarian Health and Food Safety Authority (LGL), was used for Bayesian inference and prediction at a regional scale. Information on daily number of reported infections and deaths were available up to June 30th 2020 for all 96 Bavarian districts. The relevant data used for the modelling is shown in the Supplementary Table S2. An overview of the Bavarian data can be found on the LGL homepage^29^.

## Appendix B tools to assess fat-tail properties of distributions

The *maximum-to-sum plot* relies on a consequence of the law of large numbers. For a sequence *X*_1_, *X*_2_, …, *X*_*n*_ of non-negative independently and identically distributed random variables, if *E*[*X*_p_] *<* ∞ for *p* = 1, 2, 3, …, then 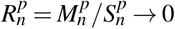 for increasing *n*. Here, 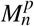 is the maximum and 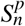 is the sum over all 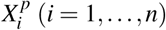. If the ratio 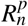 does not converge to 0 for any plotted *p*, no matter how many observations *n* are used, it proposes that no finite specific moments are likely to exist.

The *Zipf plot* is the double-logarithmic (log-log) plot of the empirical survival function. Possible fat tails can be identified in the presence of a linearly decreasing behaviour of the plotted curve. The Zipf plot shows a necessary but not sufficient condition for fat tails^30^.

A further graphical tool to detect fat-tail features is a plot of the mean excess function plot (*meplot*). If a random variable *X* is possibly fat tailed, its mean excess function *e*_*X*_ (*u*) = *E*[*X* − *u*| *X > u*] should grow linearly in the threshold *u* and should identify the actual power law tail^17^. Figure S1 shows the meplot of COVID-19 fatalities.

A useful tool for the analysis of tails is the *non-parametric Hill estimator*^17^. *For a collection of random variables X*_1_, …, *X*_*n*_ let *X*_(1)*n*_, …, *X*_(*n*)*n*_ the corresponding order statistic. The tail parameter *ξ* can be estimated as

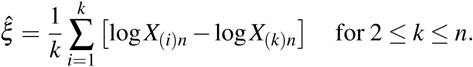

The *Hill plot* presents the estimate of 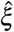 against *k*.

**Figure S1.**
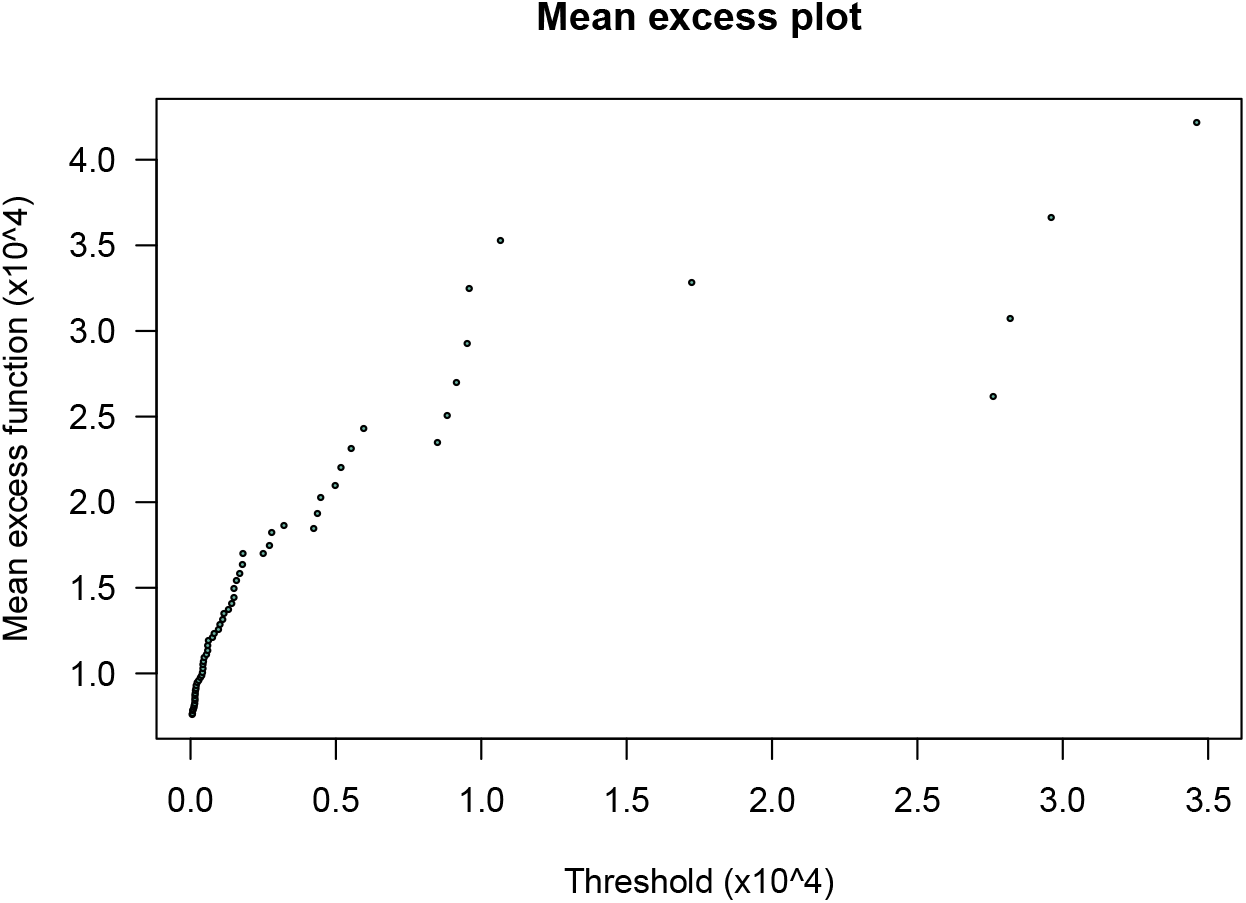
Mean excess plot of COVID-19 fatalities.

## Appendix C Bayesian prediction of peak incidences

Here we present the WinBUGS doodle used to analyze the incidence data and to provide a prediction of peak incidences during a new wave of the COVID-19 epidemic together with a short R script for the accompanying analyses. The obtained results can be saved into a text document using “coda” within WinBUGS and further processed for example in R.

### WinBUGS doodle for the Poisson model for incidences

~~~
model
{
for (i in 1:N)
{
log(mu[i])<-alpha + beta[J[i]] * (i -1 - t.change)
J[i] <- 1 + step(i -1 - t.change)
values[i]∼dpois(mu[i])
}
alpha∼dnorm(0,0.001)
for(j in 1 : 2)
{
beta[j] ∼ dnorm(0.0,1.0E-6)
}
t.change ∼ dunif(20,50)
for (k in 1:N)
{
X[k]∼dpois(mu[k])
}
Total.sum<-sum(X[])
}
Data
list(values = c(1, 2, 2, 4, 6, 6, 10, 19, 51, 20,
20, 86, 111, 119, 136, 252, 231, 169, 270, 474, 755, 822, 862,
1109, 559, 722, 1218, 1678, 1703, 1644, 1421, 861, 1039, 1841,
1989, 1941, 1859, 1496, 722, 1124, 1532, 1549, 1261, 939, 764,
419, 464, 753, 965, 864, 776, 519, 386, 373, 503, 598, 571, 531,
309, 187, 339, 362, 304, 291, 221, 137, 92, 164, 213, 269, 234,
210, 123, 78, 188, 212, 188, 208, 134, 101, 58, 87, 108, 193,
93, 117, 79, 94, 89, 134, 116, 87, 78, 83, 37, 20, 37, 54, 84,
58, 23, 37, 38, 44, 75, 35, 33, 22, 18, 35, 43, 43, 45, 40, 33,
28, 41, 45, 56, 57, 85, 42, 27, 29, 1),N=125)
Inits
list(alpha=7.6,beta=c(0.2,-0.05), t.change=35)
~~~

### WinBUGS doodle for the negative binomial model for incidences

The data and the inits are the same as in case of the Poisson model and thus not shown in this doodle.

~~~
model
{
for (i in 1:N)
{
log(r[i])<-alpha + beta[J[i]] * (i - 1 - t.change) + logit(pp)
rr[i]<-round(r[i])+1
J[i] <- 1 + step(i - 1 - t.change)
values[i]∼dnegbin(pp,rr[i])
}
alpha∼dnorm(0,0.001)
for(j in 1 : 2)
 {
beta[j] ∼ dnorm(0.0,1.0E-6)
}
t.change ∼ dunif(20,50)
for (k in 1:N)
{
X[k]∼dnegbin(pp,rr[k])
}
Total.sum<-sum(X[])
}
~~~

### Death data for WinBUGS

We used the same models as provided for the incidences. The death data included in the modelling is provided here.

~~~
Data
list(values = c(0, 0, 0, 0, 0, 0, 0, 1, 0, 0, 0,
0, 0, 0, 1, 0, 2, 1, 2, 2, 2, 9, 9, 7, 9, 13, 15, 18, 32, 29,
35, 40, 63, 61, 52, 57, 65, 70, 65, 72, 72, 86, 69, 78, 85, 59,
69, 67, 68, 74, 73, 62, 67, 54, 51, 45, 38, 40, 48, 41, 39, 38,
43, 32, 26, 19, 26, 21, 28, 20, 22, 13, 12, 19, 12, 14, 8, 19,
14, 5, 12, 10, 11, 16, 6, 12, 10, 6, 8, 7, 1, 6, 6, 5, 4, 5,
6, 4, 4, 4, 2, 2, 0, 0, 3, 2, 3, 1, 1, 3, 2, 0, 5, 1, 0, 1, 1,
0, 0, 0, 1, 0, 1, 0, 0),N=125)
~~~

### R script

~~~
rm(list=ls())
dat.1<-read.csv(“20200630_LGL_times.csv”,header=T,sep=“,”,as.is=T)
library(chron)
# dat.1 is Bavaria
dat.1$datum.new<-chron(dates=dat.1$datum,format=c(“d.m.y”))
ll.1<-nrow(dat.1)
dat.1$time<-0:(ll.1-1)
# Replace missings
# If no death or case is reported, nothing happened
dat.1$deaths.new<-ifelse(is.na(dat.1$deaths),0,dat.1$deaths)
dat.1$cases.new<-ifelse(is.na(dat.1$cases),0,dat.1$cases)
time.start<-30
dat.1.mod<-dat.1[-(1:time.start),]
dat.1.mod$time<-dat.1.mod$time-dat.1.mod$time[1]
total.ind<-sum(dat.1.mod$cases.new)
total.death<-sum(dat.1.mod$deaths.new)
tt<-dat.1.mod$time
yy<-dat.1.mod$cases.new
zz<-dat.1.mod$deaths.new
tt.rr<-tt[-(1:40)]
yy.rr<-yy[-(1:40)]
zz.rr<-zz[-(1:40)]
ww.ind<-(zz.rr!=0)
summary(lm(log(yy.rr)∼tt.rr))
summary(lm(log(zz.rr[ww.ind])∼tt.rr[ww.ind]))
coef(lm(log(yy.rr)∼tt.rr))
coef(lm(log(zz.rr[ww.ind])∼tt.rr[ww.ind]))
# prepare WinBUGS data
winbugs.cases.data<-list(time=tt,values=yy)
dput(winbugs.cases.data,”winbugs_cases.dat”)
winbugs.death.data<-list(time=tt,values=zz)
dput(winbugs.death.data,”winbugs_death.dat”)
# Entering the WinBugs Output
# there are 1000 cycles
# beta has two dimensions
# The prediction looks at 125 days
N.days<-125
# Results for the Poisson model
alpha.po<-read.table(“alpha_Poisson.txt”,header=T,sep=“\t”)
beta.po<-read.table(“beta_Poisson.txt”,header=T,sep=“\t”)
x.change.po<-read.table(“t_change_Poisson.txt”,header=T,sep=“\t”)
pred.po<-read.table(“X_Poisson.txt”,header=F,sep=“\t”)
total.sum.po<-read.table(“Tot_Sum_Poisson.txt”,header=F,sep=“\t”)
beta.po<-rbind(c(10001, 0.1838),beta.po)
# Results for the negbin model
alpha.nb<-read.table(“alpha_NegBin.txt”,header=T,sep=“\t”)
beta.nb<-read.table(“beta_NegBin.txt”,header=T,sep=“\t”)
x.change.nb<-read.table(“t_change_NegBin.txt”,header=T,sep=“\t”)
pred.nb<-read.table(“X_NegBin.txt”,header=F,sep=“\t”)
total.sum.nb<-read.table(“Tot_Sum_NegBin.txt”,header=F,sep=“\t”)
beta.nb<-rbind(c(10001,0.192),beta.nb)
# Figure WinBUGS Results
pdf(“Figure_XXX.pdf”)
par(mfrow=c(2,2))
# Panel A: Observed data, mean time courses Poisson, Negbin
plot(yy∼tt,type=“n”,log=“y”,xlab=“Time in Days since 27.02.2020”,ylab=“Number”)
# Mean time course Poisson
for (i in sample(1:1000,300,replace=T))
{
 yyy<-alpha.po[i,2]+beta.po[i,2]*(tt-x.change.po[i,2])*(1+
 sign(x.change.po[i,2]-tt))/2+ beta.po[i+1000,2]*
 (tt-x.change.po[i,2])*(1+sign(tt-x.change.po[i,2]))/2
 lines(exp(yyy)∼tt,col=“red”)
}
# Mean time course NegBin
for (i in sample(1:1000,300,replace=T))
{
 yyy<-alpha.nb[i,2]+beta.nb[i,2]*(tt-x.change.nb[i,2])*(1+
 sign(x.change.nb[i,2]-tt))/2+
 beta.nb[i+1000,2]*(tt-x.change.nb[i,2])*(1+sign(tt-x.change.nb[i,2]))/2
 lines(exp(yyy)∼tt,col=“grey”)
}
points(tt,yy,pch=20)
title(“A”,cex=10)
# Panel:B Observed data, future predictions Poisson, Negbin
plot(yy∼tt,type=“n”,log=“y”,xlab=“Time in Days since 27.02.2020”,ylab=“Number”)
mm<-matrix(pred.nb[,2],byrow=T,nrow=N.days)
for (i in sample(1:1000,300,replace=T)) lines(mm[,i]∼tt,col=“grey”)
mm<-matrix(pred.po[,2],byrow=T,nrow=N.days)
for (i in sample(1:1000,300,replace=T)) lines(mm[,i]∼tt,col=“red”)
points(tt,yy,pch=20)
title(“B”,cex=10)
# Panel: C predicted peak distribution
# Negative binomial
mm<-matrix(pred.nb[,2],byrow=T,nrow=N.days)
possible.max.values<-apply(mm,2,max)
possible.max.values.nb<-possible.max.values
dens.max.nb<-density(possible.max.values)
qq.nb<-quantile(possible.max.values.nb,probs=c(0.5,0.8,0.95,0.99))
qq.nb
# Poisson
mm<-matrix(pred.po[,2],byrow=T,nrow=N.days)
possible.max.values<-apply(mm,2,max)
possible.max.values.po<-possible.max.values
dens.max.po<-density(possible.max.values)
qq.po<-quantile(possible.max.values.po,probs=c(0.5,0.8,0.95,0.99))
qq.po
mi.x<-min(c(dens.max.nb$x,dens.max.po$x))
mi.y<-min(c(dens.max.nb$y,dens.max.po$y))
ma.x<-max(c(dens.max.nb$x,dens.max.po$x))
ma.y<-max(c(dens.max.nb$y,dens.max.po$y))
boxplot(cbind(possible.max.values.nb,possible.max.values.po),
 names=c(“Neg. Binomial”,”Poisson”),ylab=“Peak Number”)
abline(h=yy[order(yy)][120:125],lty=3)
title(“C”,cex=10)
# Panel: D predicted cumulative case distribution
# Negative binomial and Poisson
tt.nb<-total.sum.nb[,2]
tt.po<-total.sum.po[,2]
qq.nb<-quantile(tt.nb,probs=c(0.5,0.8,0.95,0.99))
qq.po<-quantile(tt.po,probs=c(0.5,0.8,0.95,0.99))
boxplot(tt.nb,tt.po,names=c(“Neg. Binomial”,”Poisson”),
ylab=“Cumulative number of cases”)
abline(h=sum(yy),lty=3)
title(“D”,cex=10)
par(mfrow=c(1,1))
dev.off()
# Days with more than 1000 incidences
mm<-matrix(pred.nb[,2],byrow=T,nrow=N.days)
# noi: number of interest
dd.nb<-apply(mm,2,function(x,noi=1000){sum(x>noi)})
table(dd.nb)
mm<-matrix(pred.po[,2],byrow=T,nrow=N.days)
# noi: number of interest
dd.po<-apply(mm,2,function(x,noi=1000){sum(x>noi)})
table(dd.po)
~~~

**Table S2.** Bavarian data of incidences and deaths between February 27th and June 30th, 2020.

| Date | Reported deaths | Reported incidences | Date | Reported deaths | Reported incidences | Date | Reported deaths | Reported incidences |
| --- | --- | --- | --- | --- | --- | --- | --- | --- |
| 27.02.20 | 0 | 1 | 09.04.20 | 69 | 1261 | 21.05.20 | 6 | 93 |
| 28.02.20 | 0 | 2 | 10.04.20 | 78 | 939 | 22.05.20 | 12 | 117 |
| 29.02.20 | 0 | 2 | 11.04.20 | 85 | 764 | 23.05.20 | 10 | 79 |
| 01.03.20 | 0 | 4 | 12.04.20 | 59 | 419 | 24.05.20 | 6 | 94 |
| 02.03.20 | 0 | 6 | 13.04.20 | 69 | 464 | 25.05.20 | 8 | 89 |
| 03.03.20 | 0 | 6 | 14.04.20 | 67 | 753 | 26.05.20 | 7 | 134 |
| 04.03.20 | 0 | 10 | 15.04.20 | 68 | 965 | 27.05.20 | 1 | 116 |
| 05.03.20 | 1 | 19 | 16.04.20 | 74 | 864 | 28.05.20 | 6 | 87 |
| 06.03.20 | 0 | 51 | 17.04.20 | 73 | 776 | 29.05.20 | 6 | 78 |
| 07.03.20 | 0 | 20 | 18.04.20 | 62 | 519 | 30.05.20 | 5 | 83 |
| 08.03.20 | 0 | 20 | 19.04.20 | 67 | 386 | 31.05.20 | 4 | 37 |
| 09.03.20 | 0 | 86 | 20.04.20 | 54 | 373 | 01.06.20 | 5 | 20 |
| 10.03.20 | 0 | 111 | 21.04.20 | 51 | 503 | 02.06.20 | 6 | 37 |
| 11.03.20 | 0 | 119 | 22.04.20 | 45 | 598 | 03.06.20 | 4 | 54 |
| 12.03.20 | 1 | 136 | 23.04.20 | 38 | 571 | 04.06.20 | 4 | 84 |
| 13.03.20 | 0 | 252 | 24.04.20 | 40 | 531 | 05.06.20 | 4 | 58 |
| 14.03.20 | 2 | 231 | 25.04.20 | 48 | 309 | 06.06.20 | 2 | 23 |
| 15.03.20 | 1 | 169 | 26.04.20 | 41 | 187 | 07.06.20 | 2 | 37 |
| 16.03.20 | 2 | 270 | 27.04.20 | 39 | 339 | 08.06.20 | 0 | 38 |
| 17.03.20 | 2 | 474 | 28.04.20 | 38 | 362 | 09.06.20 | 0 | 44 |
| 18.03.20 | 2 | 755 | 29.04.20 | 43 | 304 | 10.06.20 | 3 | 75 |
| 19.03.20 | 9 | 822 | 30.04.20 | 32 | 291 | 11.06.20 | 2 | 35 |
| 20.03.20 | 9 | 862 | 01.05.20 | 26 | 221 | 12.06.20 | 3 | 33 |
| 21.03.20 | 7 | 1109 | 02.05.20 | 19 | 137 | 13.06.20 | 1 | 22 |
| 22.03.20 | 9 | 559 | 03.05.20 | 26 | 92 | 14.06.20 | 1 | 18 |
| 23.03.20 | 13 | 722 | 04.05.20 | 21 | 164 | 15.06.20 | 3 | 35 |
| 24.03.20 | 15 | 1218 | 05.05.20 | 28 | 213 | 16.06.20 | 2 | 43 |
| 25.03.20 | 18 | 1678 | 06.05.20 | 20 | 269 | 17.06.20 | 0 | 43 |
| 26.03.20 | 32 | 1703 | 07.05.20 | 22 | 234 | 18.06.20 | 5 | 45 |
| 27.03.20 | 29 | 1644 | 08.05.20 | 13 | 210 | 19.06.20 | 1 | 40 |
| 28.03.20 | 35 | 1421 | 09.05.20 | 12 | 123 | 20.06.20 | 0 | 33 |
| 29.03.20 | 40 | 861 | 10.05.20 | 19 | 78 | 21.06.20 | 1 | 28 |
| 30.03.20 | 63 | 1039 | 11.05.20 | 12 | 188 | 22.06.20 | 1 | 41 |
| 31.03.20 | 61 | 1841 | 12.05.20 | 14 | 212 | 23.06.20 | 0 | 45 |
| 01.04.20 | 52 | 1989 | 13.05.20 | 8 | 188 | 24.06.20 | 0 | 56 |
| 02.04.20 | 57 | 1941 | 14.05.20 | 19 | 208 | 25.06.20 | 0 | 57 |
| 03.04.20 | 65 | 1859 | 15.05.20 | 14 | 134 | 26.06.20 | 1 | 85 |
| 04.04.20 | 70 | 1496 | 16.05.20 | 5 | 101 | 27.06.20 | 0 | 42 |
| 05.04.20 | 65 | 722 | 17.05.20 | 12 | 58 | 28.06.20 | 1 | 27 |
| 06.04.20 | 72 | 1124 | 18.05.20 | 10 | 87 | 29.06.20 | 0 | 29 |
| 07.04.20 | 72 | 1532 | 19.05.20 | 11 | 108 | 30.06.20 | 0 | 1 |
| 08.04.20 | 86 | 1549 | 20.05.20 | 16 | 193 |  |  |  |

## Notes

### Competing Interest Statement

The authors have declared no competing interest.

### Funding Statement

No external funding was received.

